# Multi-omics Integration Identifies Genes Influencing Traits Associated with Cardiovascular Risks: The Long Life Family Study

**DOI:** 10.1101/2024.03.04.24303657

**Authors:** Sandeep Acharya, Shu Liao, Wooseok J. Jung, Yu S. Kang, Vaha A. Moghaddam, Mary Feitosa, Mary Wojczynski, Shiow Lin, Jason A. Anema, Karen Schwander, Jeff O Connell, Mike Province, Michael R. Brent

## Abstract

The Long Life Family Study (LLFS) enrolled 4,953 participants in 539 pedigrees displaying exceptional longevity. To identify genetic mechanisms that affect cardiovascular risks in the LLFS population, we developed a multi-omics integration pipeline and applied it to 11 traits associated with cardiovascular risks. Using our pipeline, we aggregated gene-level statistics from rare-variant analysis, GWAS, and gene expression-trait association by Correlated Meta-Analysis (CMA). Across all traits, CMA identified 64 significant genes after Bonferroni correction (p ≤ 2.8×10^−7^), 29 of which replicated in the Framingham Heart Study (FHS) cohort. Notably, 20 of the 29 replicated genes do not have a previously known trait-associated variant in the GWAS Catalog within 50 kb. Thirteen modules in Protein-Protein Interaction (PPI) networks are significantly enriched in genes with low meta-analysis p-values for at least one trait, three of which are replicated in the FHS cohort. The functional annotation of genes in these modules showed a significant over-representation of trait-related biological processes including sterol transport, protein-lipid complex remodeling, and immune response regulation. Among major findings, our results suggest a role of triglyceride-associated and mast-cell functional genes *FCER1A, MS4A2, GATA2, HDC*, and *HRH4* in atherosclerosis risks. Our findings also suggest that lower expression of *ATG2A*, a gene we found to be associated with BMI, may be both a cause and consequence of obesity. Finally, our results suggest that *ENPP3* may play an intermediary role in triglyceride-induced inflammation. Our pipeline is freely available and implemented in the Nextflow workflow language, making it easily runnable on any compute platform (https://nf-co.re/omicsgenetraitassociation).

## Introduction

The Long Life Family Study (LLFS) is a multi-center, longitudinal family study that enrolled families enriched for exceptional longevity to discover genetic, behavioral, and environmental factors contributing to healthy aging and long life. LLFS enrolled 4,953 participants in 539 families, including probands, offspring, grandchildren, and spouses. Participants are primarily of European ancestry (99%). The data it has generated include microarray genotypes, whole genome sequences, gene expression from whole blood, and biomarkers of health and aging. Healthy aging and long life are heritable traits [1, 2] and the LLFS cohort is exceptional in both [3]. The LLFS probands and offspring were less likely to have diabetes, chronic pulmonary disease, and peripheral artery disease than participants in the Cardiovascular Health Study (CHS) and Framingham Heart Study (FHS) in the same age group [4]. High-density cholesterol levels were higher, and pulse pressure and triglycerides were lower in the LLFS cohort than in CHS and FHS [4]. In this work, we look for genes that affect cardiovascular health in the LLFS population and the biological processes through which they work. We focus on 11 traits associated with cardiovascular risks spanning four categories: pulmonary (forced expiratory volume, forced vital capacity, and the ratio of the two), lipids (high-density lipoprotein, low-density lipoprotein, triglycerides, total cholesterol), anthropometric (BMI, BMI-adjusted waist), and cardiovascular (pulse, ankle-brachial index) [5-9].

Genome-wide association studies (GWAS) have identified many loci for cardiovascular-related domains, including pulmonary function [10, 11], lipids [12], obesity and body fat distribution [13, 14], and blood pressure and ankle-brachial index [15, 16]. However, GWAS has some well-known limitations. Testing millions of individual variants requires extremely small p-values and hence very large cohorts. When GWAS does identify statistically significant variants, it is difficult to determine which are causal and which are merely tagging a causal variant in linkage disequilibrium [17]. If a causal non-coding variant is found, it is often unclear which gene it acts through. We set out to address these challenges. To reduce the multiple testing burden, we aggregated variant-level GWAS p-values for common variants (minor allele frequency (MAF) > 5%) to obtain gene-level p-values, used a SKAT-based [18] analysis method [19] to calculate gene-level p-values for rare variants (MAF < 5%), [18-21] and calculated gene-level p-values for association between measured gene expression levels and traits (Transcriptome-wide association studies (TWAS); throughout this paper, TWAS refers to association with measured gene expression levels, not predicted levels). We combined the gene-level p-values from TWAS, GWAS, and rare variant analysis (RVA) using a meta-analysis approach that accounts for expected correlations among these [22]. Aggregating variants to the gene level creates strong evidence about which gene is implicated, which can be difficult when focusing on individual variants. By incorporating evidence from TWAS, we reduce the chance that a significant gene is simply tagging a nearby gene in LD (since LD does not induce correlation in the expression levels of nearby genes). TWAS alone has a different problem – gene expression may be associated with a trait because it is affected by the trait, rather than affecting the trait, or by a confounding factor affecting both trait and gene expression. However, when there is supporting evidence from genetic variants, that is less likely.

To further investigate a gene’s potential for causally affecting a trait, we started with the hypothesis that, among genes statistically associated with a trait, the most likely to be causal are those that interact with other statistically associated genes (1) through a common molecular system, and (2) serve a common biological function. To identify genes that interact with other statistically associated genes through a common molecular system, we searched for network modules in protein-protein interaction networks whose genes, as a group, are significantly enriched for genes with suggestive/significant p-values from correlated meta-analysis. To identify common biological functions served by module genes, we looked for GO biological process terms significantly overrepresented among genes in the enriched modules [23].

This paper makes three contributions. First, it presents 64 genes from meta-analysis that are genome-wide significant for at least one of 11 traits associated with cardiovascular risks, of which 29 are replicated in the FHS population. Second, it presents 13 protein-protein interaction network modules significantly enriched in genes with comparatively low meta-analysis p-values for at least one of the traits. Three such modules are replicated in the FHS population. Third, it presents software that researchers can use to conduct similar analyses. The software is packaged as a Nextflow pipeline, which containerizes each analysis step, simplifies the maintenance of software dependencies, and enables deployment across multiple computing environments, including cloud computing provided by data repositories [24]. The software pipeline and complete documentation can be found at https://nf-co.re/omicsgenetraitassociation/. Figure 1 depicts the pipeline.

**Fig 1:**
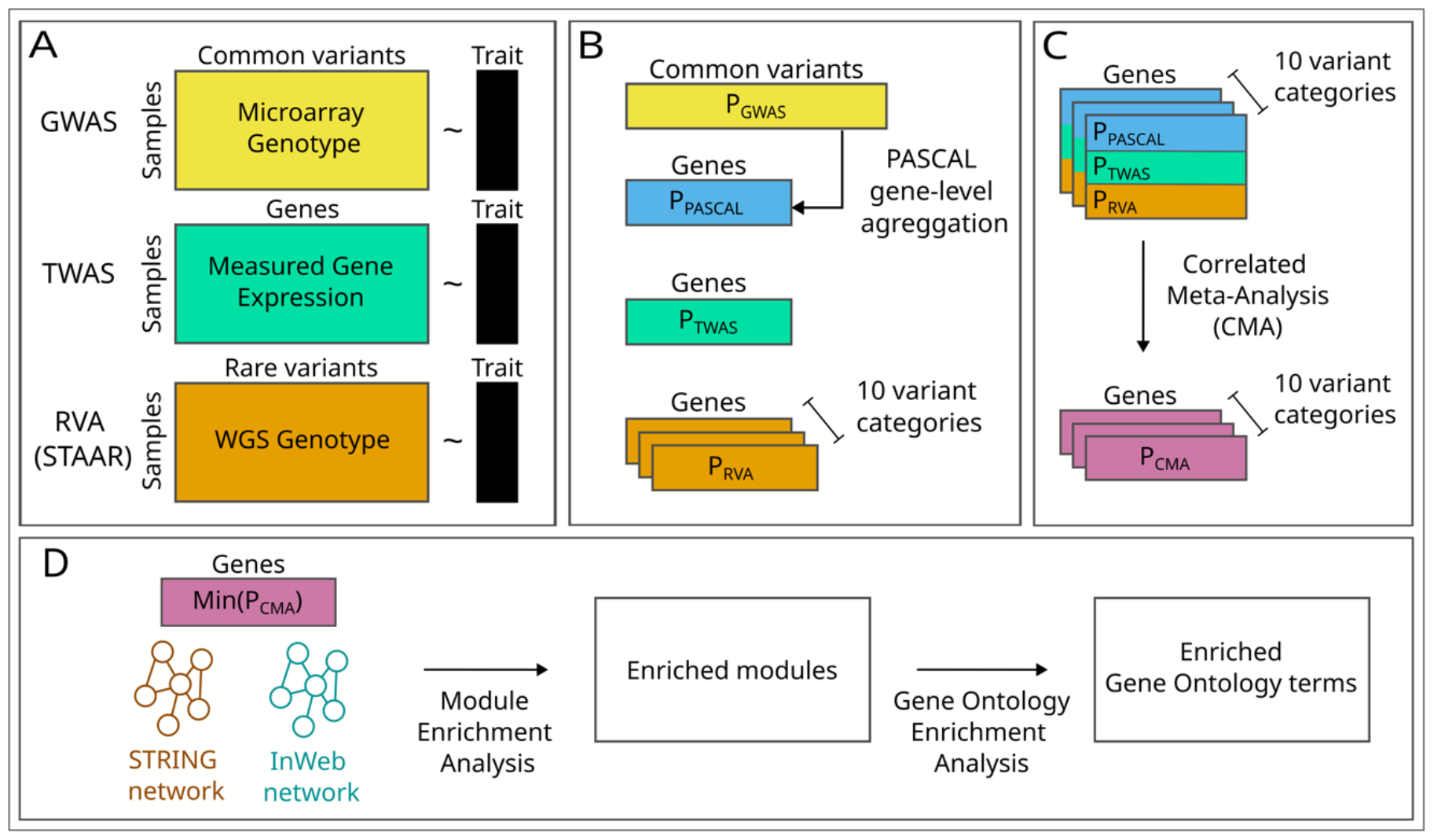
Pipeline diagram. A) Inputs to GWAS, TWAS, and RVA. B) GWAS output is fed into PASCAL, which calculates gene-level p-values. TWAS outputs gene-level p-values. STAAR splits variants into ten functional categories and outputs 10 p-values per gene. C) Correlated meta-analysis (CMA) is run 10 times. Each run uses outputs from PASCAL and TWAS together with one variant category of STAAR, outputting 10 p-values. D) For each gene, the minimum p-value from 10 CMA runs is fed into module enrichment analysis, which is also performed by PASCAL. PASCAL outputs enriched modules and their p-values. E) Gene ontology over-representation analysis identifies biological processes with significant over-representation among genes in each module.

## Material and methods

### Participants

The recruitment procedure, eligibility criteria, and enrollment of the LLFS participants have been previously described [4]. We used data from the first clinical exam, which started in 2006 and recruited 4953 individuals from 539 families. Across 11 studied traits, the participants ranged from *n* = 2528 to 4166 for GWAS, *n* = 595 to 1200 for gene expression level–trait association, and *n* = 2528 to 4166 for rare-variant analysis. Descriptive statistics for all the traits and covariates can be found in File S3. The number of participants in each analysis depended on the number of participants with data for the trait, microarray genotypes for GWAS, whole genome sequencing for rare-variant analysis, and RNA-Seq for TWAS.

### Cardiovascular-related traits

We used trait values from the first clinical exam. BMI was calculated as weight (kg)/height (m)^2^, and waist as the average of three abdominal circumference measurements in cm. Pulse was calculated as the average of three measurements of the sitting pulse. FEV1 and FVC were measured in a portable spirometer (EasyOne, NDD Medical Technologies, Andover, MA), as previously reported [4]. High-density lipoprotein (HDL), low-density lipoprotein (LDL), triglyceride (TG), and total cholesterol (TC) were assessed and analyzed by the LLFS central laboratory based at the University of Minnesota, as previously reported [4]. Participants were excluded if fasting < 8 hours for LDL, TG, and TC. Ankle-brachial index (ABI) was derived as the average of the right and left ankle-arm blood pressure ratio. We excluded participants with non-compressible arteries (ABI >= 1.4). For all analyses, each of the traits was adjusted for age, sex, field center, and square of the age. Waist and pulse were additionally adjusted for BMI. FEV1, FVC, and FEV1/FVC were adjusted for height and smoking. LDL and TC were adjusted for statin use, and TG was log-transformed. All traits were also adjusted for the top 10 genetic principal components stepwise. After covariate adjustments, all traits were inverse normal transformed.

### GWAS and gene level aggregation of GWAS results

GWAS SNP-chip data for the LLFS participants were produced using Illumina 2.5 million HumanOmni array. Genotypes were called using Bead Studio. SNPs were removed if their call rate was less than 98%, if their allele frequency in the LLFS population was < 1% or > 99%, if they had an allelic mismatch with 1000 Genomes Project (1000Gp3v5), or if they displayed excess heterozygosity relative to Hardy Weinburg Equilibrium (p < 1E-6). A single-SNP association test was done for all SNPs passing the quality filter by using a linear mixed model. Family relatedness was accounted for using a pedigree-based kinship matrix, and an additive genetic model was assumed. The SNP-level summary statistics from GWAS for SNPs with minor allele frequency >= 5 % were input to PASCAL[25]. The SNPs were assigned to a gene if they lied within 50kb of the gene body. PASCAL uses the sum of the chi-squared approach to calculate a gene-level p-value. Document S1 describes the GWAS and gene level aggregation process for the FHS population.

### Gene-expression to trait association (TWAS)

The RNA extraction and sequencing were carried out by the McDonnell Genome Institute at Washington University (MGI). Total RNA was extracted from PAXgene™ Blood RNA tubes using the Qiagen PreAnalytiX PAXgene Blood miRNA Kit (Qiagen, Valencia, CA). The Qiagen QIAcube extraction robot performed the extraction according to the company’s protocol. The RNASeq data were processed with the nf-core/RNASeq pipeline version 3.3 using STAR/RSEM and otherwise default settings (https://zenodo.org/records/5146005). RNASeq on whole blood samples from the LLFS participants in the first clinical visit was used for the analysis. Genes with less than three counts per million in greater than 98.5% of samples were filtered out from the analysis. Samples with greater than 8% of reads in intergenic regions were also filtered out. The resulting set were transformed using DESeq’s [26] variance stabilizing transform (VST) function. The VST transformed gene expression levels were adjusted for base covariates: age, age squared, sex, field center, percent of reads mapping to intergenic sequence, and the counts of red blood cells, white blood cells, platelets, monocytes, and neutrophils. The gene expression level was also adjusted stepwise for the RNA-seq batch and the top 10 principal components of gene expression. For each trait, the adjusted gene expression residuals were used as a predictor, and the adjusted trait was used as a response variable in a linear mixed model implemented in MMAP [27]. A kinship matrix generated by MMAP from the LLFS pedigree was used to account for family relatedness. For traits with genomic inflation factor (GIF) > 1.1, the p-values were adjusted using BACON [28]. The same RNA-Seq processing steps were implemented for replication in the FHS dataset.

### Rare-variant analysis (RVA) using STAAR

LLFS Whole Genome Sequence (WGS) was produced by MGI using 150bp Illumina reads. Variant calls with read depth less than 20 or greater than 300 were set to missing. Variants with call rate < 90% and those with excess heterozygosity (p < 1E-6) were excluded from the analysis. Missing genotype calls in the LLFS cohort were filled in using the call with the highest phred-scale likelihood from GATK. Bi-allelic SNVs with MAF < 5% and passing the above quality filters were input to STAAR [19] for variant set association tests using SKAT [18]. We also employed burden testing [29-32] and Aggregate Cauchy Association Test (ACAT) [33, 34] as implemented in the STAAR framework. However, the resulting p-value distributions from these tests displayed a U-shaped pattern, deviating from the expected uniform distribution under the null hypothesis so we did not use them.

For each gene, variants are split into 10 functional categories, and an omnibus association test is performed for each category for each gene weighted by functional annotations from the FAVOR database [35], which is curated by the TOPMed Consortium. The 10 functional categories include synonymous, missense, putative loss of function (plof), promoter CAGE, promoter DHS, enhancer CAGE, enhancer DHS, upstream, downstream, and untranslated region (UTR) [19, 21, 35]. A minimum of 2 variants is required in a category to perform a SKAT test. Document S1 describes the WGS data processing steps for the FHS population.

### Correlated Meta-analysis (CMA)

CMA [22] combined gene p-values from GWAS (after aggregation by PASCAL), TWAS, and RVA while preventing Type I errors by accounting for dependencies between individual analyses under the null as described [22, 36]. GWAS, TWAS, and RVA were performed on overlapping individuals from LLFS’s first clinical visit. Furthermore, genetic variants affect gene expression. Therefore, each pair of inputs to CMA may be correlated. Since STAAR outputs 10 p-values per gene, one for each category, we ran CMA 10 times resulting in 10 p-values per gene.

### Module enrichment analysis and Gene Ontology (GO) Over-representation Analysis

We started with modules (highly connected subnetworks) from two protein-protein interaction (PPI) networks, the STRING functional PPI network [37] and the InWeb physical PPI network [38] which were identified by the best-performing methods in a DREAM challenge [39]: random walk algorithm R1 for STRING and modularity optimization algorithm M2 for InWeb. These modules and the gene-level p-values were input to PASCAL’s module enrichment algorithm [25]. Genes with p-values from fewer than two CMA input sources were removed from the modules. The module enrichment p-values from PASCAL were corrected for the total number of modules tested using Bonferroni correction. GO over-representation analysis was done on the set of genes in each enriched module by using WebGestaltR package (version: 0.4.6,) with the following configuration: (organism: hsapiens, method: ORA, enrichDatabase: GO Biological Process, FDRMethod: BH, FDRThreshold = 0.05) [23]. The affinity propagation feature in WebGestaltR was used to eliminate GO biological processes with highly overlapping member genes.

### Framingham Heart Study (FHS) replication

FHS is a multi-generational study to identify genetic and environmental factors affecting cardiovascular and other diseases [40, 41]. We used the data on the FHS participants from grandchildren and offspring spouse generation who attended examination 2 for replication purposes [40, 41]. Across 11 studied traits, the participants ranged from *n* = 2512 to 3341 for GWAS, *n* = 1080 to 1380 for TWAS, and *n* = 921 to 1233 for rare-variant analysis. Descriptive statistics for all the traits and covariates can be found in File S3. We use the same pipeline described in Fig 1 to replicate the LLFS results in the FHS population. Replication analysis was done on genes that were significant in LLFS by CMA or by any of the CMA inputs: TWAS, GWAS, or RVA. For each trait, a gene is replicated if it meets the Bonferroni significance threshold, which is adjusted for the number of genes that were significant in the LLFS population in GWAS and TWAS, or for the number of gene-category pairs of significant genes in CMA and STAAR. The significance threshold used for GWAS, TWAS, RVA, and CMA for both LLFS and FHS can be found in Table S3. A module is replicated if it is significantly enriched after applying Bonferroni correction based on the number of significantly enriched modules across all traits in the LLFS population.

### GWAS Catalog Search

We used NHGRI-EBI GWAS Catalog database (version: v1.0.2-associations_e109) [42] to check if the gene-trait associations with suggestive/significant signals from GWAS, TWAS, RVA, and CMA have a previously known trait-associated genome-wide significant variant within the 50 kb region of the gene body. Genes matching this criterion are designated as “previously associated in GWAS Catalog” throughout the paper. It is important to note that the presence of previously known trait-associated variants in a 50kb region around the trait-associated gene’s body does not necessarily establish a causal role for the gene on the trait. However, we use this broad criterion to ensure that we classify genes with any hint of previous implication as “previously associated,” minimizing the risk of incorrectly classifying them as novel findings.

## Results

Figure 1 shows the flowchart of the multi-omics integration pipeline we used to identify genes and biological processes affecting 11 cardiovascular-related traits. We implemented it as a Nextflow workflow, which containerizes each process [24]. This greatly simplifies the maintenance of software dependencies and enables easy deployment across various computing environments. The complete pipeline documentation can be found at https://nf-co.re/omicsgenetraitassociation/.

### Gene-level aggregation of GWAS

File S3 shows the characteristics of study participants for covariates and 11 cardiovascular-related traits for GWAS, TWAS, and RVA. We employed GWAS on all traits. Genomic inflation factors (GIFs) for all traits (Table S1) indicate no systematic inflation, technical bias, or population stratification. We then aggregated GWAS summary statistics to the gene level using PASCAL [25]. After aggregation, GIFs range from = 1.07 to 1.21 (GIFs: Table S2 correction, 30 gene-trait associations were genome-wide significant across five traits – low-density lipoprotein (LDL, 9 genes), total cholesterol (TC, 7 genes), High-density lipoprotein (HDL, 4 genes), waist (1 gene), and triglycerides (TG, 9 genes). 26 of these gene-trait pairs are previously associated in GWAS Catalog [42]. We replicated 9/30 genome-wide significant gene-trait associations in the FHS population using aggregated GWAS (Table S3). One of those genes for TG, *BUD13-DT* (p = 2.25 × 10^−8^), is not previously associated in GWAS Catalog. However, *BUD13-DT* is a divergent transcript and shares 82 of the 83 genetic variants that are aggregated to the gene level with *BUD13. BUD13* is previously associated in GWAS Catalog.

#### Transcriptome-wide Association Study (TWAS)

We conducted TWAS on the 11 traits. After using BACON [28] to correct for inflation when GIF > 1.10, the GIFs range from 1.01 to 1.16 (GIFs: Table S2). After Bonferroni correction, 77 gene-trait associations were genome-wide significant across five traits – TC (5), BMI (21), HDL (21), FVC (1), and TG (29) (Table S5). 9 of the 77 genes are previously associated in GWAS Catalog, and 57 of the 77 associations were replicated in the FHS population (Table S5, Table S10). The direction of the effect matches between the LLFS and the FHS population for all 57 FHS-replicated associations. Of 21 genes significant for HDL, 18 were also significant for TG. Consistent with the inverse relationship between HDL and TG traits, the HDL and TG β-values had opposite signs for all 18 genes. 50 of the 57 replicated gene-trait associations are not previously associated with the corresponding traits in the GWAS Catalog [42]. Among 9 genes previously associated in GWAS Catalog, 7 were replicated in FHS – *HCAR3* (BMI p = 1.12 × 10^−8^), *HCK* (BMI p = 3.91 × 10^−7^), *SLC45A3* (HDL p = 1.42 × 10^−15^), *LINC02458* (HDL p = 7.17 × 10^−15^), *ABCG1* (HDL p = 1.98 × 10^−9^), *ENPP3* (HDL p = 9.68 × 10^−9^), and *ABCA1* (TG p = 5.22 × 10^−9^). These previously associated genes in GWAS Catalog have GWAS Catalog reported trait-associated variant(s) within the 50 kb region of the gene body. The genome-wide significance of these previously associated genes after TWAS and their replication in the FHS population suggests a potential role as mediators linking trait-associated variants to traits. For 3 of the 7 replicated TWAS genes with trait-associated variants within 50 Kb, the variants are assigned to other, closer genes in the GWAS Catalog. Our analysis suggests the following reassignments: rs3747973 from *NUCKS1* and *Metazoa_SRP* to *SLC45A3*, rs2245133 from *MED23* to *ENPP3*, rs2245611 from *HCAR1* and *DENR* to *HCAR3*, and rs6489191 from *KNTC1* and *HCAR2 to HCAR3* [42].

#### Rare variant analysis (RVA)

We applied RVA on the same 11 traits using the STAAR package [19]. STAAR splits variants into 10 functional categories and performs 10 variant set tests for each gene. The GIFs of all 110 STAAR-category-trait combinations (10 categories by 11 traits) range from 0.77 to 1.20 (GIFs: Table S2) After Bonferroni correction, we identified 194 unique gene-trait associations at the genome-wide significant levels for ABI (13), LDL (2), TC (1), BMI (16), FEV1 (24), FEV1/FVC (8), HDL (7), FVC (49), TG (2), pulse (3), and waist (69) (Table S6). 22/194 are previously associated genes, and 5/194 associations were replicated in the FHS population (Table S6, Table S10). *OR52A1* (p = 2.56 × 10^−8^) is genome-wide significant for ABI, was replicated in FHS, and not previously associated in GWAS Catalog [42]. The low replication rate in FHS may stem from LLFS’s unique cohort enriched for exceptional longevity. Rare variants unique to LLFS could drive the phenotype under study. One example is *NABP1* (p = 2.12 × 10^−8^), which is genome-wide significant for HDL in the upstream category. Two rare variants upstream of this gene (rs10931513, rs10177406) have minor allele counts of 5 and are present in the same group of individuals. GWAS on these variants for HDL shows that each one individually has a suggestive p-value (betas = 2.02, p < 8 × 10^−6^). Other genes that are significant in LLFS but not replicated in FHS warrant further investigation.

#### Correlated meta-analysis (CMA)

After aggregating gene-level p-values from PASCAL, TWAS, and each category of RVA [22], we obtained 10 category-specific p-values for each gene. The GIFs across all 110 category-trait combinations ranged from 0.98 to 1.29 (GIFs: Table S2). After Bonferroni correction, we identified 64 significant genes across 9 traits – LDL (6), TC (1), BMI (4), FEV1 (3), FEV1FVC (1), HDL (15), FVC (8), TG (23), and waist (3), of which 21 are previously associated genes (Table S7). Twenty-nine of 64 gene-trait associations were replicated in the FHS population, of which 9 genes are previously associated in the GWAS Catalog [42] (Table S7, Table S10). We identified 20 genes that were not previously associated and were replicated in the FHS population (Table 1), including 14 for TG, 5 for HDL, and 1 for BMI.

**Table 1:** Genes that are genome-wide significant after CMA and FHS-replicated. 20/29 genes have not been previously associated with the trait in the GWAS Catalog. *TOMM40* for TC, *ATG2A* for BMI, and *APOC3* for TG are more significant after CMA than after PASCAL, TWAS, and RVA alone. *TOMM40* for LDL, *CETP* for HDL, and *APOC3, LPL*, and *SLC45A3* for TG have suggestive or genome-wide significant p-values from more than one analysis. The remaining genes are CMA-significant due to a highly significant p-value in one analysis.

| Genes | Trait | CMA p-value | GWAS catalog? | PASCAL p-value | TWAS p-value | RVA p-value | RVA category |
| --- | --- | --- | --- | --- | --- | --- | --- |
| *TOMM40* | LDL | 4.9E-17 | Yes | 1.9E-12 | 7.9E-01 | 1.8E-17 | synonymous |
| *TOMM40* | TC | 2.7E-11 | Yes | 1.2E-06 | 2.8E-01 | 2.3E-10 | synonymous |
| *ATG2A* | BMI | 6.1E-08 | No | 1.8E-01 | 1.8E-07 | 7.5E-04 | downstream |
| *AKAP12* | HDL | 1.3E-11 | No | 1.0E-01 | 3.1E-16 | NA | NA |
| *CETP* |  | 1.6E-18 | Yes | 9.5E-22 | 8.8E-01 | 1.3E-11 | missense |
| *CPA3* |  | 3.4E-10 | No | 2.6E-01 | 6.2E-16 | NA | NA |
| *FCER1A* |  | 2.2E-08 | No | 6.6E-01 | 2.8E-16 | NA | NA |
| *GATA2* |  | 2.3E-11 | No | 5.5E-01 | 3.9E-21 | NA | NA |
| *HERPUD1* |  | 1.0E-11 | Yes | 8.7E-17 | 1.2E-01 | NA | NA |
| *LINC02458* |  | 1.0E-08 | Yes | 4.3E-01 | 7.2E-15 | NA | NA |
| *MS4A2* |  | 5.5E-11 | No | 1.5E-01 | 6.7E-16 | NA | NA |
| *SLC45A3* |  | 1.8E-12 | Yes | 7.9E-02 | 1.4E-15 | 1.4E-03 | enhancer_DHS |
| *AKAP12* | TG | 2.9E-27 | No | 4.7E-01 | 4.7E-53 | NA | NA |
| *APOA5* |  | 7.3E-08 | Yes | 2.2E-09 | NA | 4.5E-02 | promoter_CAGE |
| *APOC3* |  | 2.1E-11 | Yes | 8.0E-09 | NA | 1.0E-04 | plof_ds |
| *CA8* |  | 8.4E-11 | No | 3.8E-01 | 1.0E-20 | 4.8E-02 | enhancer_CAGE |
| *CPA3* |  | 3.6E-28 | No | 2.5E-01 | 8.2E-51 | NA | NA |
| *ENPP3* |  | 4.5E-13 | No | 1.5E-01 | 1.8E-26 | 1.7E-01 | promoter_DHS |
| *FCER1A* |  | 1.7E-18 | No | 8.6E-01 | 1.1E-41 | NA | NA |
| *GATA2* |  | 6.2E-35 | No | 3.4E-01 | 4.9E-66 | NA | NA |
| *GCSAML* |  | 9.8E-15 | No | 4.8E-01 | 7.9E-28 | NA | NA |
| *HDC* |  | 2.4E-28 | No | 3.7E-01 | 4.2E-67 | 4.4E-02 | synonymous |
| *HRH4* |  | 6.6E-10 | No | 3.7E-01 | 1.6E-16 | 2.0E-02 | missense |
| *LINC02458* |  | 3.6E-18 | No | 6.3E-01 | 6.5E-37 | NA | NA |
| *LPL* |  | 5.0E-08 | Yes | 4.0E-08 | 2.7E-01 | 5.4E-04 | missense |
| *MS4A2* |  | 3.1E-34 | No | 7.0E-03 | 2.2E-50 | NA | NA |
| *MS4A3* |  | 4.9E-18 | No | 6.8E-03 | 7.1E-23 | NA | NA |
| *NTRK1* |  | 2.3E-07 | No | 2.0E-01 | 1.0E-10 | NA | NA |
| *SLC45A3* |  | 3.1E-30 | No | 1.9E-01 | 7.5E-56 | 6.5E-05 | enhancer_DHS |

CMA accounts for the correlation between p-values from PASCAL, TWAS, and RVA outputs, but we observed minimal correlation between the TWAS output and PASCAL or RVA output. The absolute median tetrachoric correlation across all trait-category pairs ranges from 0.001 to 0.009 for [TWAS, RVA] and from 0.004 to 0.017 for [TWAS, PASCAL]. The absolute median correlation across trait-category pairs is slightly higher between RVA and PASCAL, ranging from 0.02 to 0.05 (Table S8).

The p-values from CMA were inputted to PASCAL’s module enrichment analysis method, which identifies modules whose genes, as a group, have significantly lower p-values than would be expected by chance after Benjamin-Hochberg correction for the number of tested modules [25]. We used modules from the InWeb (physical) [38] and STRING (functional) [37] protein-protein interaction (PPI) networks. We identified 13 enriched modules across 7 traits – LDL (1), TC (1), BMI (3), FEV1FVC (1), HDL (2), FVC (2), and TG (3) (Table 2). Of the 13, 6 modules are in the physical network and 7 are in the functional one. Three of 13 modules were replicated in FHS (see Methods). One replicated STRING module (cma-STRING-104) is enriched for genes with suggestive/significant p-values for both HDL and for TG. It contains three genome-wide significant TG genes – *APOA5* (p = 1.56 × 10^−15^), *APOC3* (p = 2.08 × 10^−11^), and *APOA4* (p = 1.56 × 10^−15^), of which *APOA5* and *APOC3* were replicated in FHS. The module also contains two genome-wide significant HDL genes - *APOC3* (p = 1.64 × 10^−8^) and *CETP* (p = 1.61 × 10^−18^), of which *CETP* was replicated (Fig 2a). Functional annotation of genes in this module showed a significant over-representation of multiple biological processes. Notably, the top 5 most significant biological processes are lipid-related – sterol transport, glycerolipid catabolism, protein-lipid complex remodeling, phospholipid transport, and plasma lipoprotein particle assembly (Table S9). Another FHS-replicated STRING module for TG contains two replicated genes for TG that are not previously associated in GWAS Catalog – *MS4A2* (p = 3.14 × 10^−34^) and *FCER1A* (p = 1.71 × 10^−18^) along with *SYK* (not suggestive or significant) and *CBL* (suggestive). However, the expression level of *MS4A2* and *FCER1A* has been associated with TG in two previous studies [43, 44]. The over-represented biological processes for this module include immune-related processes such as positive regulation of immune response, mast-cell degranulation, and T-cell-activation (Fig 2b). The most significant biological processes for 7 other significantly enriched lipid-related modules – LDL (1), TC (1), HDL (1), BMI (3), and TG (1), are primarily lipid-related or immune-related processes (Table 2). The genes in enriched modules and over-represented biological processes for enriched modules can be found in File S1.

**Table 2:** 13 modules that are significantly enriched for genes with low CMA p-values. cma-InWeb-46 is enriched for both LDL and TC. cma-STRING-104 is enriched for HDL and TG and is FHS-replicated. The most significant Gene Ontology (GO) biological processes across lipid traits are primarily lipid-related or immune-response-related.

| Enriched Module | Trait | Bonf. corrected p-value | FHS-Replicated | Most Significant Biological Process (BP) | FDR (GO BP) |
| --- | --- | --- | --- | --- | --- |
| cma-STRING-2 | BMI | 4.9E-02 | No | T Cell Activation | 0 |
| cma-InWeb-5 | BMI | 1.3E-02 | No | innate immune response activating signal transduction | 1.1E-05 |
| cma-InWeb-7 | BMI | 9.6E-03 | No | immune response regulating signaling pathway | 0 |
| cma-InWeb-58 | FEV1FVC | 4.4E-03 | No | regulation of canonical Wnt signaling pathway | 0 |
| cma-STRING-11 | FVC | 4.6E-02 | No | Retinol metabolic process | 2.4E-12 |
| cma-STRING-188 | FVC | 3.3E-02 | No | JAK STAT cascade | 1.8E-11 |
| cma-InWeb-7 | HDL | 1.5E-02 | No | positive regulation of immune response | 0 |
| cma-STRING-104 | HDL | 3.5E-07 | Yes | sterol transport | 0 |
| cma-InWeb-46 | LDL | 6.3E-04 | No | protein lipid complex remodeling | 4.9E-10 |
| cma-InWeb-46 | TC | 4.8E-03 | No | plasma lipoprotein particle remodeling | 4.9E-10 |
| cma-STRING-104 | TG | 6.7E-04 | Yes | sterol transport | 0 |
| cma-STRING-193 | TG | 2.0E-05 | Yes | positive regulation of immune response | 0 |
| cma-STRING-48 | TG | 4.0E-02 | No | establishment of protein localization to organelle | 0 |

**Fig 2:**
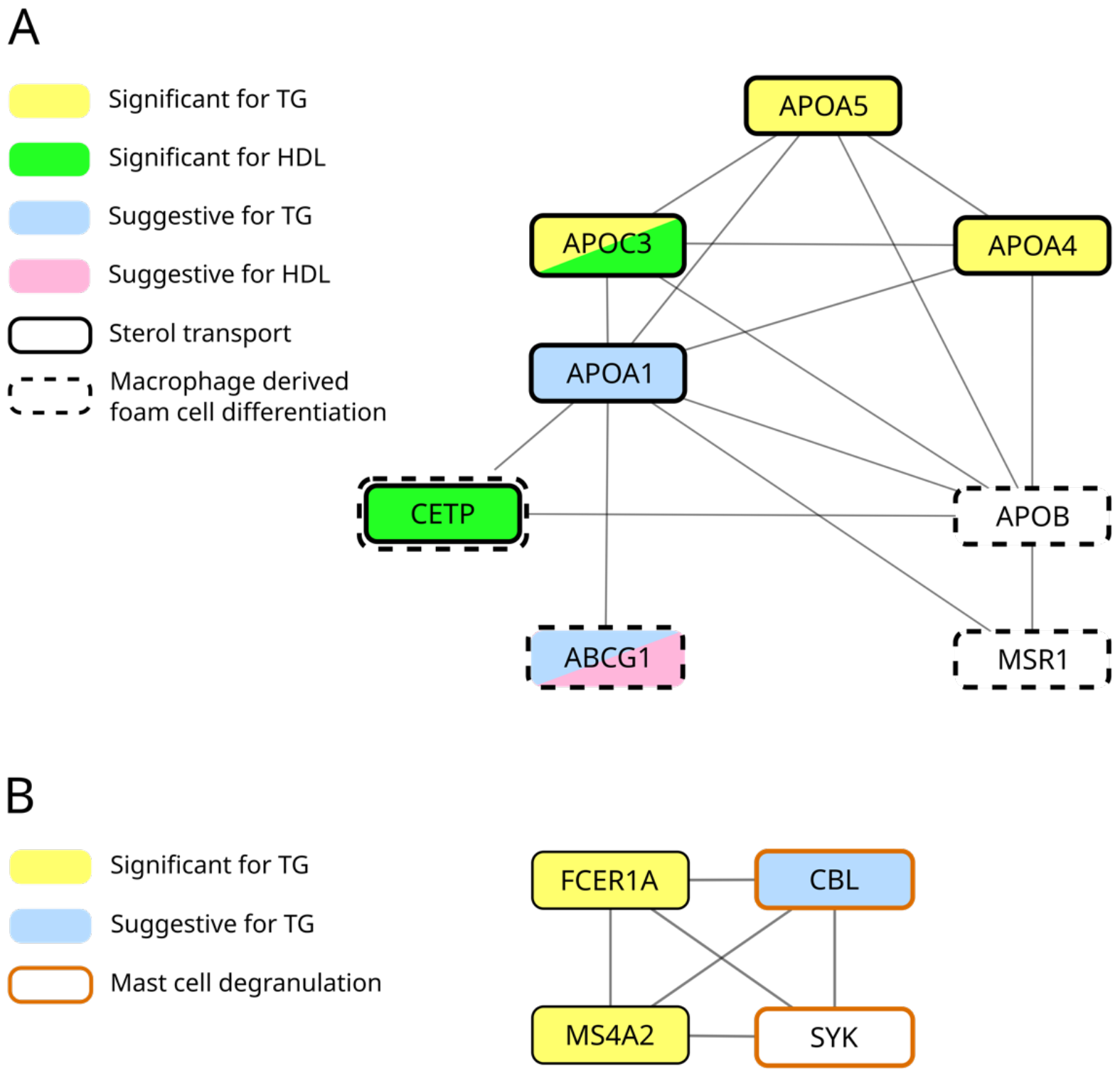
Sub-modules within enriched modules. Genes with *P* < 10^−4^ are annotated as suggestive. Module enrichment analysis identifies trait-related genes missed by association analysis. (A) Module cma-STRING-104 is enriched for both TG and HDL. *APOB* is not significant for TG but directly interacts with genes with suggestive or significant p-values for TG. *APOB* and *MSR1* participate in macrophage-derived foam cell differentiation with two genome-wide significant genes (*CETP* and *ABCG1)*. (B) *SYK* is not genome-wide significant after CMA. *SYK* interacts with significant genes for TG, *FCER1A* and *MS4A2*, and participates in mast-cell degranulation.

## Discussion

### The value of correlated meta-analysis

Using correlated meta-analysis (CMA), we developed a strategy to integrate evidence from GWAS, TWAS, and rare-variant analysis (RVA). The summary statistics from all four analyses can be found in File S2. After CMA, we identified 64 genome-wide significant genes across 9 cardiovascular-related traits. Of 29 CMA-significant and FHS-replicated genes, *TOMM40* for TC, *ATG2A* for BMI, and *APOC3* for TG (triglycerides) are more significant after CMA than in PASCAL, TWAS, and RVA alone (Table 1). *TOMM40* for LDL, *CETP* for HDL, and *APOC3, LPL*, and *SLC45A3* for TG have suggestive or genome-wide significant p-values from more than one input analysis (Table 1). The rest of the genes have strong evidence from TWAS. Modestly significant genes with support from only one of GWAS, TWAS, or RVA were filtered out by CMA.

Prior work using meta-analysis has primarily focused on integrating evidence from multiple GWAS on different cohorts [45-47] or identifying shared/pleiotropic genetic effects across multiple traits [36, 48]. Our approach integrates evidence from GWAS, TWAS, and RVA. Wang et al. 2020 [49] performed a meta-analysis similar to ours by integrating methylation data (EWAS), TWAS, and GWAS gene-level statistics. However, the TWAS and EWAS statistics came from a single cohort without replication and the meta-analysis also did not account for the correlation between EWAS, TWAS, and GWAS statistics from the same cohort. These are expected to be correlated because genetic variants and methylation both affect gene expression [49].

Within the individual analyses, the replication rate in the FHS population for RVA (5/194) is lower than for GWAS (9/30) or TWAS (57/77). The low replication rate for RVA is expected because the FHS sub-population with whole genome sequencing and measured traits and covariates is much smaller than the LLFS cohort (File S3). For the most significant functional categories of the 194 RVA-significant genes in LLFS, only ∼4% (59 / 1530) of the variants are present and analyzed in FHS.

### *ATG2A* and its link to obesity

Autophagy-Related Protein 2 Homolog A (*ATG2A)* is a genome-wide significant gene (p = 6.11 × 10^−8^) for BMI after CMA and is replicated in FHS (p= 1.5 × 10^−4^). *ATG2A* is not previously associated in the GWAS Catalog. The ATG2A protein plays a role in autophagosome formation, regulation of lipid droplet morphology, and lipid-droplet dispersion during autophagy [50, 51]. In vitro experiments have shown that low expression of ATG2A can disrupt normal autophagy. Velikkakath et al. reported that silencing *ATG2A/ATG2B* via siRNA in HeLa cells leads to the aggregation of large lipid droplets [50]. *ATG2A/ATG2B* double knockout in HEK293 cells led to an incomplete autophagy process [51]. The association between the expression level of *ATG2A* and BMI is genome-wide significant with a negative beta coefficient, which means higher expression of *ATG2A* is associated with lower BMI (beta = -0.7, *P* = 1.8 × 10^−7^). Obesity increases the inhibition of autophagy [52], so the lower expression of *ATG2A*, a pro-autophagy gene, may be a consequence of high BMI. On the other hand, increasing autophagy by genetic or pharmacological mechanisms protects mice from obesity and sequelae such as insulin resistance and fatty liver [52], so higher expression of *ATG2A* may protect against obesity and consequent cardiovascular risk [53]. Indeed, autophagy regulation has been proposed as a therapy to reduce the risk of obesity-associated cardiovascular diseases [54]. Thus, *ATG2A* may participate in a positive feedback loop in which lower expression of *ATG2A* is both a cause and a consequence of obesity.

### *ENPP3*: a potential mediator of TG-induced inflammation

*ENPP3* is genome-wide significant (p = 4.52 × 10^−13^) for TG after CMA and replicated in FHS (p *=* 1.41 × 10^−11^). It encodes ecto-nucleotide pyrophosphatase-phosphodiesterase 3, one of several enzymes that hydrolyze extracellular ATP and thereby tamp down chronic inflammation [55]. Extracellular ATP is a powerful “alarmin” that signals cellular damage, activates immune cells, and causes inflammation [55], a key element of atherosclerosis [56]. ENPP3-/- mouse cells exhibit lower ATP hydrolysis compared to WT cells [57]. The expression leathervel of *ENPP3* is genome-wide significant for TG with a negative coefficient (beta = -1.04, *P =* 1.75 × 10^−26^) and the direction of effect is the same in FHS. The expression level of *ENPP3* has been previously associated with TG in two prior studies with the same direction of effect [43, 44]. A 2023 bidirectional Mendelian randomization study found a significant effect of TG on *ENPP3* expression but no evidence for reverse causation [43]. Thus, reduced expression of *ENPP3* and subsequent increase in extracellular ATP concentration may be one of the mechanisms by which high TG induces inflammation and promotes atherosclerosis [56].

### Role of mast cell functional genes in atherosclerosis risks

*FCER1A, MS4A2, GATA2, HDC*, and *HRH4* are genome-wide significant for TG in LLFS CMA and replicated in FHS. None of them has a TG-associated variant within 50k in the GWAS Catalog. All 5 genes play a role in either mast-cell activation, mast-cell proliferation, or secretion of pro-inflammatory markers [58-65]. Active mast cells affect atherosclerosis risks. In mice, local activation of adventitial mast cells during atherogenesis increases plaque size, macrophage apoptosis, vascular leakage, and intraplaque hemorrhage [66]. *FCER1A* and *HDC* have also been experimentally linked to atherosclerosis. Homozygous deletion of FCER1A reduced atherosclerosis in Apoe -/- mice [61]. Similarly, HDC-/- mice exhibited reduced atherosclerotic lesions in an Apoe-/- background [63]. Using Mendelian randomization, Dekkers et al. found a significant effect of TG on all 5 genes but no evidence for reverse causation [43]. This is consistent with the fact that elevated TG causes inflammation [67] and that these genes are pro-inflammatory. Surprisingly, the association between TG and the expression of these pro-inflammatory genes is not positive, as would be expected based on the inflammatory effect of high TG levels. In fact, we see a significant negative association for all 5 genes (Table 1). The expression level of these genes has been previously associated with TG in two different studies with the same direction of effect [43, 44]. One explanation for the lack of a positive correlation between TG and these pro-inflammatory genes is that we have measured gene expression in whole blood, whereas inflammation associated with atherosclerosis occurs in plaques. However, the existence of such a strong and consistent negative correlation between TG and the expression of these genes is an intriguing mystery that demands further experimental investigation.

Module and GO enrichment analysis identified an additional gene, *SYK*, which may affect atherosclerosis risk via a similar mechanism. *SYK* lies in an enriched TG-module (cma-STRING-193) in which genes involved in mast-cell degranulation are significantly overrepresented (Fig 2b). *SYK* directly interacts with *FCER1A* and *MS4A2*, two genes with known mast cell functions [58-60]. An experimental study has shown that treating mice with *SYK* inhibitors significantly reduced atherosclerosis lesions in atherosclerosis-prone mice [68]. This suggests that combined module and GO analysis can identify important trait-related genes that are not genome-wide significant.

### A flexible and easy-to-ease pipeline

We introduced a multi-omics integration pipeline (Fig. 1) and provided a NextFlow implementation that is easily run on a wide variety of platforms, from laptops to large compute clusters (https://nf-co.re/omicsgenetraitassociation/). While we used our multi-omics integration approach to aggregate signals from GWAS, TWAS, or RVA, our pipeline can also take in gene-level summary statistics from epigenome-wide association studies (EWAS) [69]. While we used modules from the STRING and InWeb PPI networks, our pipeline can also take in modules from other networks, such as those linking transcription factors to their target genes. This flexibility makes the pipeline useful for a wide range of research problems.

In the future, we plan to enhance the pipeline to address some limitations. Currently, we aggregate variant-level statistics from GWAS to the gene level based on proximity to the gene. This could be improved by aggregating variants in the genes’ regulatory regions using publicly available resources on regulatory regions and their target genes [70, 71]. The current meta-analysis approach does not offer weighted aggregation of different input sources. This could be improved by providing options to use various meta-analysis tools. The current implementation offers only STAAR for rare variant analysis. This could be improved by offering other, less complex options.

## Data Availability

The code used to do all association analyses is available at https://nf-co.re/omicsgenetraitassociation/. The summary results from GWAS, TWAS, RVA, and CMA on all 11 traits for the LLFS cohort are available in File S2. The input datasets have not been deposited in public repositories due to data use constraints.

## Acknowledgments

We are grateful to the entire Long Life consortium, its participants, and its investigators, without whom this work would not have been possible. We would particularly like to Dr. Bharat Thyagarajan, Dr. Allison Kuipers, and Hannah Campbell for consultations on the 11 cardiovascular risk traits analyzed here. This work was supported grant AG063893 from the National Institute on Aging.

The Framingham Heart Study is conducted and supported by the National Heart, Lung, and Blood Institute (NHLBI) in collaboration with Boston University (Contract No. N01-HC-25195, HHSN268201500001I and 75N92019D00031). This manuscript was not prepared in collaboration with investigators of the Framingham Heart Study and does not necessarily reflect the opinions or views of the Framingham Heart Study, Boston University, or NHLBI.

