## Supplementary material for "Multi-omics Integration Identifies Genes Influencing Traits Associated with Cardiovascular Risks: The Long Life Family Study": Document S1

**Supplemental Material and Methods**

**Rare Variant Analysis (RVA) – FHS Cohort**

WGS (freeze 9b) data was used in RVA. Bi-allelic SNVs that pass all QC filters were kept, resulting in ~52 million variants. Samples were excluded if they were either sequence controls, not sequenced in blood, or had FREEMIX percentage > 3%. Additionally, samples were removed if they had a mean depth of < 30x or < 95% of sites covered at 10x or < 80% at 20x. Bi-allelic SNVs with MAF < 5% and passing the above quality filters were inputted into STAAR [1] for variant set association tests using SKAT [2]. STAAR is a variant set association test for uncommon and rare variants. For each gene, variants are split into 10 functional categories, and an omnibus association test is performed for each category for each gene weighted by functional annotations from the FAVOR database [3], which is curated by the PubMed Consortium. The 10 functional categories include synonymous, missense, putative loss of function (plof), promoter CAGE, promoter DHS, enhancer CAGE, enhancer DHS, upstream, downstream, and untranslated region (UTR). A minimum of 2 variants is required in each category to perform a SKAT test.

**GWAS Analysis – FHS Cohort**

The Affymetrix 550K array (Affymetrix 500K mapping array plus Affymetrix 50K supplemental array) in NCBI36 was first lifted over to GRCh37 using Picard [4, 5]. Variant and sample-level quality control were performed using PLINK1.9 [6]. SNPs with call rate < 97% and HWE p < 1E-6 were excluded. SNPs were also excluded if they ranked among the top 0.2% SNPs with the highest Mendel errors in their MAF bin (bin width of 0.05). Individual genotype calls were set to be missing in all families where a Mendel error occurred if an SNP did not reach this threshold. Samples with call rate < 97%, mismatched sex, and excess Mendel errors were excluded. The quality-controlled genotypes were then prepared for the imputation [7] and uploaded to the Michigan Imputation Server [8]. Genotypes were phased with Eagle v2.4 [9] and then imputed to the HRC1.1 European panel [10] using Minimac4 [11]. Bi-allelic SNPs with imputation R2 >= 0.8, MAF > 5%, and exist in WGS were selected and merged with WGS genotypes. Cohen’s Kappa [12, 13] was used to check concordance between WGS and the imputed genotype, and only SNPs with high concordance (kappa >= 0.998) were kept. WGS genotypes were preferentially used when samples had both WGS and imputed genotypes. This set of 1,290,650 SNPs was lifted to GRCh38 and used for GWAS. A single SNP association testing was done for all SNPs passing the quality filter using a mixed linear model regression. Family relatedness was accounted for using a pedigree-based kinship matrix, and an additive genetic model was assumed. The SNP-level summary statistics from GWAS were used as an input to the PASCAL[14] for gene-level aggregation of summary statistics. The SNPs are assigned to a gene if they are within 50kb of the gene body. The sum of the chi-squared approach described in PASCAL[14] was used to calculate the gene-level p-value for each gene.
